# Population mortality before and during armed conflict in Yemen: geospatial and statistical analysis of cemetery data

**DOI:** 10.1101/2023.08.29.23294777

**Authors:** Francesco Checchi, Emilie Koum Besson, Ola Ali, Mervat Alhaffar, Naji Saeed, Yolanda Vasquez, Terri Freemantle, Momin Ashraf, Chris Reeve, Colin Scott, Timothy Lingard, Andy Norris

## Abstract

Since 2014, Yemen is affected by crisis conditions due to armed conflict. Evidence on the impact of this large-scale crisis on mortality is lacking. We analysed archive very high-resolution satellite imagery from a sample of Yemeni subdistricts to quantify changes in burial incidence attributable to the crisis.

We identified possible cemeteries through remote and ground sources in 24 sampled subdistricts. After initial triage and extensive steps to improve the interpretability of archive imagery spanning the period 2011 to 2021, a pool of crowd workers, supervised by expert analysts and aided by an automated algorithm, annotated surface area and grave counts in sequential images from a set of analysis-eligible cemeteries. We complemented these longitudinal observations with data on different predictors including three crisis proxies (incidence of insecurity events, price of staple cereal, internal displacement), and fitted statistical models to compare predicted burials under observed and assumed counterfactual (no crisis) conditions.

We identified 561 potential cemeteries within 24 sampled subdistricts, but excluded most due to inability to geolocate them or see the cemetery and/or graves in available imagery, yielding an effective sample of 110 image observations across 35 cemeteries in 10 subdistricts. Burial rate generally decreased between 2014-2018 and rose sharply thereafter. Alternative regression models suggested that most cemeteries would have experienced lower burial rate under non-crisis conditions, with a crisis to non-crisis ratio of about two overall. The incidence of insecurity events appeared positively associated with burial rate.

This unprecedented-scale geospatial analysis of cemeteries suggests an increase in burial rates attributable to crisis conditions in a non-representative, disproportionately urban sample of Yemen. The study identifies key challenges of such an analysis. We discuss possible methodological ways forward to further explore the feasibility and validity of this option for mortality estimation in settings with insufficient vital events registration and limited ground access.

## Introduction

Since late 2014, Yemen has been affected by armed conflict resulting in worsened food security, internal displacement and disruption of public services [1]. The country has also been separated into areas controlled by opposing authorities. The crisis in Yemen has been described as the world’s largest, but the extent to which it has affected population mortality across the country remains unclear, depriving response actors and other stakeholders of critical evidence for benchmarking the war’s impacts and the adequacy of response efforts [2]. While a credible estimate of people killed by war injuries (about 110,000 as of end 2022 [3]) has been established based on media and civil society report monitoring, the death toll indirectly attributable to the conflict could be substantial, as suggested by the occurrence over the past few years of repeated phases of food insecurity [4, 5], reduced health service functionality [6, 7], forced displacement [8] and large-scale epidemics [9]. Against this backdrop, the COVID-19 pandemic would plausibly have resulted in a further mortality increase [10–12].

In Yemen, as in many low- and middle-income countries [13, 14], vital events registration is not robust, necessitating collection of alternative mortality data. In 2019, a United Nations report [15] based on scenario modelling projected that 166,000 conflict deaths and 316,000 deaths indirectly attributable to the crisis would occur by 2022, based on a median scenario. In nine purposively selected communities within Aden and Ta’iz governorates, we combined lists generated by key informants with capture-recapture statistics to estimate adult death rates, suggesting a pattern of considerably elevated mortality during the war period, compared to baseline [16]. However, this study was limited by underreporting of child deaths and the purposive nature of the sample. After the first phase of the pandemic, we also analysed sequential very high-resolution (VHR) satellite images of cemeteries in the city of Aden to estimate pandemic-attributable mortality, likely the first instance of this method’s use [17]. Here, we expand methods and scale of this geospatial data-driven approach to explore how the crisis has affected mortality patterns elsewhere in Yemen.

## Methods

### Study population and period

Our study period was January 2011 to December 2021 (11y). The current armed conflict escalated in June 2014 (start of the ‘crisis period’ in our analysis), while the first confirmed SARS-CoV-2 infection was reported on 10 April 2020, with limited testing data suggesting two distinct waves (May to September 2020 and March to May 2021) [18]: we thus consider April 2020 and beyond as the ‘pandemic period’.

Yemen is divided into 22 governorates, 335 districts and 2149 subdistricts. The latter had a mean population of 13,000 based on United Nations projections available in early 2020. We originally intended to estimate mortality based on a representative sample of Yemeni person-time: we thus selected 24 subdistricts through systematic random sampling based on probability proportional to population size along the sampling frame of all sub-districts, sorted by (i) cumulative rate of war injury deaths as reported by ACLED [19] (https://acleddata.com/data-export-tool/) and (ii) population density (as a proxy of urbanisation). We sought to collect longitudinal data on incident burials within each cemetery in each sampled subdistrict. Assuming all decedents were buried in recognisable cemeteries (i.e. burial rate = death rate), this sample size should have provided 80% power to detect an increase in the crude (all-age, all-cause) death rate of ≥ 10% from the UN-projected baseline for 2010-2015 (6 per 1000 per year), with 95% significance, design effect of 3.5 and 20% attrition. As the study progressed, however, it became clear that resource and imagery limitations precluded analysis of all cemeteries, constraining analysis to a smaller sample within which robust data could be generated (see below).

## Data sources and collection

### Lists and locations of potential cemeteries

Before undertaking cemetery identification in each of the sampled subdistricts, we searched the grey and peer-reviewed literature non-systematically and interacted with a network of Yemeni researchers and civil society members formed during a previous research project [16] to explore burial customs and how these may have changed during the war and pandemic. These information sources indicated that the vast majority of Yemenis are buried in recognised cemeteries.

We composed a longlist of potential cemeteries by (i) asking our network to liaise with contacts in each subdistrict; (ii) visually inspecting freely available VHR imagery and annotation layers within the OpenStreetMap (https://www.openstreetmap.org/), Wikimapia (https://wikimapia.org), Google Earth (https://earth.google.com/web/) and Google Maps (https://www.google.com/maps) applications, guided by pilot work in Aden [17] and Mogadishu, Somalia [20]; (iii) leveraging a professional network of Yemeni geographers, who were able to either personally geolocate cemeteries or supply information (e.g. village, nearby landmarks) to aid remote geolocation. UK- and Yemen-based geographers worked together to resolve missing coordinates of individual cemeteries, identify duplicates across the three above sources and exclude cemeteries that were not within the subdistrict boundaries or (rarely) whose surface area mostly fell outside the subdistrict.

### Satellite imagery

At the time of this study, only one satellite imagery provider (Maxar) offered imagery with the resolution (< 50 cm) required for burial identification. We purchased from Maxar all commercially available, archive VHR images that covered successfully geolocated candidate cemeteries and were acquired during the analysis period with a spatial resolution between ∼31cm to <50 cm per pixel. Images were supplied by SecureWatch (https://www.maxar.com/products/securewatch) as Ortho Natural Colour images that have been, pre-processed, pan-sharpened and corrected for illumination and geometric distortion.

### Predictor data

We used multivariate predictive models to estimate the evolution of burial rate in each cemetery, and generate counterfactual burial rate levels in the absence of a crisis (see Statistical Analysis). Some predictor variables for these models came from the geospatial analysis itself, and included image-level (whether infilling had occurred since the previous image; image quality score; see Geospatial Analysis) and cemetery-level (under northern or southern government control; urban versus rural setting) characteristics. Externally sourced predictors included (i) a geospatial dataset of Yemen’s road network [21], which we transformed into road density (Km per Km^2^ area); (ii) a crowd-sourced dataset of health facilities [22], which we combined with reconstructed population denominators [8] to estimate health facility density per 100,000 people; (iii) governorate-level estimates of under 5y mortality per 1000 live births as per the 2013 Demographic and Health Survey [23], as a proxy of baseline burial rate; (iv) georeferenced data on insecurity events and fatalities collected by the Armed Conflict Location and Event Data Project (ACLED) [19] since 2015 through intensive media monitoring and civil society reports [3]; (v) the proportion of internally displaced persons (IDPs) within the subdistrict, per month, estimated separately [8]; (vi) exposure to COVID-19, expressed as the proportion of the inter-image period within each cemetery that fell within the pandemic period; and (vii) the price of wheat, a key staple and thus proxy of food insecurity, as collected monthly in four urban markets (Aden, Al Hudaydah, Sana’a, Sa’ada) by the World Food Programme [24] (we paired each subdistrict with one of these markets based on proximity and expressed price per 2011 USD adjusted for inflation rates). For time-varying predictors (insecurity events, wheat price), we calculated the mean value during each inter-image period.

## Geospatial analysis

### Image inspection and initial shortlisting of cemeteries

Initial inspection was conducted for all cemeteries by no fewer than two experienced geospatial analysts. Potential cemeteries for which boundaries and/or graves were not visible were excluded. Remaining cemeteries were further shortlisted based on expected ease of demarcating graves and cemetery area from surrounding terrain given soil type (agricultural, rocky, sand, vegetation), vegetation cover and available imagery. Only cemeteries for which, according to the analysts, there was a realistic chance of obtaining data after image enhancement (see below) were taken forward to the next stage of analysis.

### Enhancement and quality scoring of images for shortlisted cemeteries

Given available resources, we sought to include between two (below which burial rate was unobservable) and three images per cemetery, with no more than one image per analysis subperiod (pre-crisis, crisis and pandemic). To select the best images, we firstly enhanced each available image using the `scikit-image` Python package, in three sequential steps: (i) stripping any alpha channels from the image, leaving only the UInt-8 RGB (red, green and blue) channels (if an image had only one channel, we converted it to a greyscale RGB); (ii) Contrast Limited Adaptive Histogram Equalization [25, 26] on the image, with a clip limit of 3% and a kernel size of 1/8th of the image; (iii) merging this contrast-adjusted image with the source image, with an alpha of 30% (i.e. 30% coming from the adjusted image, 70% from the source); and (iv) performing unsharp masking [27] to sharpen the image without unduly exacerbating noise, with a radius of 10 pixels, amplification (amount) of 0.5 and treating each colour channel independently. The net result of these transformations is exemplified in S1 Appendix (Fig S1).

Next, we employed crowd sourcing to score from 0 (i.e. perfect visibility) to 3 (unusably poor) each enhanced image *i* according to two dimensions of quality: (i) ‘area clarity’ or *q*_*a*,*i*_ (ability to discern the cemetery’s boundaries and any rows or blocks of graves) and (ii) ‘grave clarity’ or *q*_*g*,*i*_ (ability to identify individual graves): example imagery is shown in S1 Appendix (Fig S2, Fig S3). We relied on a crowd worker pool (primarily North American [28] and remunerated based on United States living wage, but with no geographical or educational attainment restriction other than English language comprehension) provided by Amazon’s Mechanical Turk platform, which has previously supported geospatial projects. Altogether, 1105 crowd workers contributed analysis, though we omitted instances of inadequate output. Images submitted to the crowd worker pool were accompanied by extensive help text and examples and seeded with hidden assessments to remove workers that might have been misunderstanding or gaming the task (further detail available on request). As mean *q*_*a*,*i*_ and *q*_*g*,*i*_(computed from > 5000 individual scores of 932 images) were highly correlated (S1 Appendix, Fig S4), we combined both into a single quality score 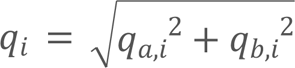. Notably, image date was not a correlate of quality (data not shown).

### Final selection of images for analysis

For each cemetery and subperiod, we focussed on the two images with best (lowest) *q*_*i*_ and excluded any whose quality was so poor that we could not identify and place point markers on graves, plus any sets of images for a given cemetery across which we could not consistently identify at least three static, ground-level features (e.g. a tree) to support tie-point orthorectification (see below). After applying these further criteria, cemeteries that had < 2 analysable images over the entire analysis period were additionally excluded. One of the cemeteries in Aden governorate had been analysed in depth as part of the previous study [17]: as geospatial methods were compatible, we included data from this cemetery.

### Surface area estimation for included images

To achieve orthorectification of images from a given cemetery (i.e. comparable geometries despite varying satellite angles), we employed the same crowd worker pool to place at least four and up to six tie-points at consistently identifiable features (e.g. graves, buildings, trees) that appeared in each image: we then used the `scikit-image` package to fit affine transformations to these tie-points; these transformations generally resulted in negligible (a few pixels) errors on cross-validation. Three tie-points would have been sufficient for the affine transformation used here, but extra tie-points allowed for a measure of error and to help identify where transformations were more likely to fail.

Within each cemetery, and starting with the highest-quality image, we then relied on a subset of highly performing crowd workers to annotate the vertices of the cemetery’s borders: the resulting polygon was used as a starting basis to annotate the previous and next images in the cemetery’s timeline, and so forth until a surface area polygon was drawn for all images (S1 Appendix, Fig S5, Fig S6). Lastly, we converted polygons into m^2^ by translating each to the orthorectified frame of the oldest image from each cemetery, using this image’s affine transformation to convert to EPSG:4326 coordinates, and then to the National Snow and Ice Data Center (NSIDC) EASE-Grid 2.0 North coordinate system (EPSG:6931, https://epsg.io/6931 [29]).

Some cemeteries were particularly sparse, with graves dotted across the entire area (S1 Appendix, Fig S7). In this instance, a polygon was placed around the exterior boundary of the site, but without computing surface area.

### Grave enumeration for included images

We attempted to help crowd workers in this task by automatically identifying and annotating likely graves through an algorithm optimised through manual inspection. The algorithm combined edge detection and peak identification techniques and relied on hyperparameters that were only weakly tailored to each individual cemetery, so as to make it applicable across all sites. More information on the algorithm’s development is available from the authors. The algorithm was tuned to prefer false negatives to false positives, as analysts found it easier to add rather than remove grave markers; despite this, it occasionally resulted in moderate sensitivity *and* specificity, as exemplified in S1 Appendix, Fig S9. The algorithm was not used for sparse cemeteries (see above).

After automatic annotation, we divided images into small tiles projected to contain no more than ≈ 100 graves in the densest of sites, assuming a minimum surface area per grave of 1.4m^2^. We then allocated tiles to crowd workers and asked them to revise and improve automated annotation by manually moving, adding or deleting algorithm-generated markers; workers also had the option to start from a blank slate by removing all automated markers, though most chose to retain and amend them. Here too, we used hidden assessments and extensive validation of crowd worker output by expert analysts to filter out unacceptable work and reduce the annotator pool to a trusted team. We used agglomerative clustering in `scikit-learn` to merge different annotators’ grave markers by assuming that markers up to within 7 pixels (∼3.5m) of each other could identify the same grave.

Lastly, so as to mitigate the problem of graves disappearing or becoming less visible over time (see below), we mapped graves forwards in time from the oldest to the most recent image, such that any graves missed in more recent images would be identified from older ones. Because orthorectification does not yield perfectly overlapping images, we resolved the position of each grave over successive images by minimising the sum of pixel distances from every grave in an older image to every grave within the same 10-pixel radius in the more recent image (this was done in Python through the `scipy.optimize.linear_sum_assignment` function), manually checking that output was acceptable. The end result is exemplified in Fig 1.

**Fig 1.**
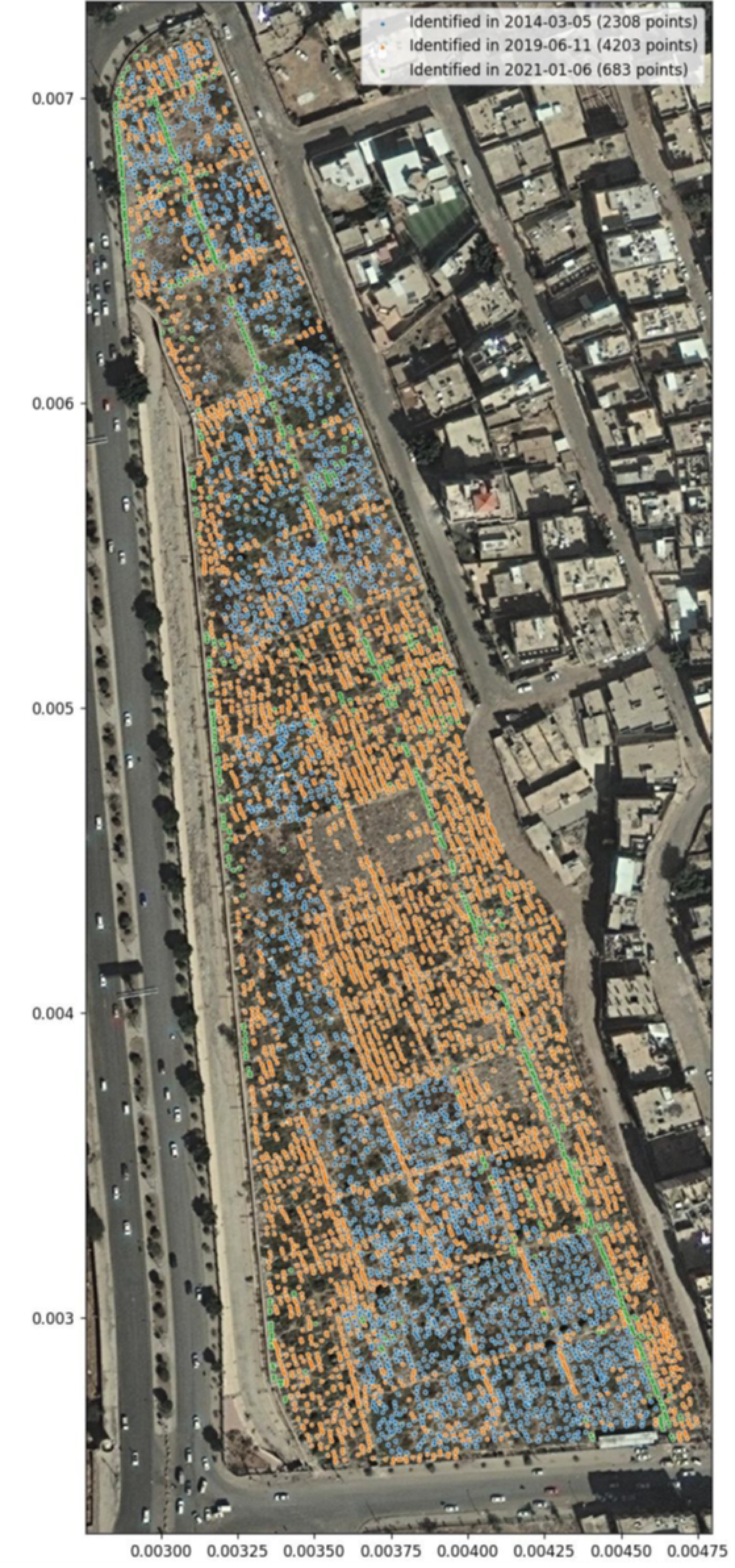
Example of a cemetery at three different time points: graves (coloured in blue) from the earliest time point are carried over and supplemented by newer graves from the next time point (coloured in orange) and the latest time point (in green). Over time, walkways separating different cemetery blocks are filled in with graves, presumably due to space running out. Satellite image © 2021 Maxar Technologies.

### Issues encountered during geospatial analysis

The above steps ran up against different recurring issues with the potential to introduce random error or bias into the analysis: these are listed in Table 1, along with data management decisions taken to address each.

**Table 1.** Issues encountered during geospatial analysis, and corresponding data management decisions.

| Issue | Potential effect | Decision |
| --- | --- | --- |
| **1. Higher quality of recent images reveals previously undetected graves.** For some cemeteries (particularly urban), increased image fidelity over time allowed identification of graves where previously not enough information had been available. | **Overestimation of burial rate** due to an artefactual increase in grave counts over time in cemetery sections already containing graves, an increase partly or fully attributable to improved signal, rather than infilling†. | Ignore grave count in all but the highest-quality image for the cemetery. Impute these missing grave counts from surface area, where possible (see Statistical Analysis). |
| **2. Serious degradation of image quality over time.** This made it hard to measure surface area changes, detect new graves or even perform orthorectification of older images. | **Underestimation of burial rate**, or at least potential for high inaccuracy. | Omit observations from these images from analysis. |
| **3. Vegetation growth** covers more recent images, partly impeding identification of new graves or surface area expansion. | **Underestimation of burial rate** due to some sections of the cemetery becoming invisible. | Case-by-case inspection of images in question: treat observations from these images as missing if it is clear that infilling† or area expansion are occurring in nonvisible sections of the cemetery. |
| **4. Sections of cemeteries with very different grave sizes**, possibly constituting separate areas for adults and children / infants. Smaller graves may be too small to annotate. | **Minor data loss.** Grave counts could not be done for one cemetery. Other instances of this issue did not impede grave annotation. | Omit all observations from this single cemetery. |
| **5. Poor tie-point correction**, leading to inaccuracy in orthorectification (i.e. successive images aligning imperfectly) due to bad choice of tie-points or underlying topography. | **Negligible** under- or overestimation of burial rate. This issue was largely mitigated through re-analysis by expert analysts. | Ignore issue. |
| **6. No change in surface area or grave counts is seen over time,** suggesting the cemetery may have been closed to new burials during the entire analysis period. | **Uninformative observations.** Data cannot be used to estimate the burial rate. | Omit cemetery from analysis (i.e. retain only cemeteries that were ever active during the analysis period). |

† We use the term 'infilling' to denote the frequently encountered practice of digging new graves within cemetery sections Potential cemeteries for which boundaries that are already in use, e.g. by using walkways in between graves or any other empty space.

## Statistical analysis

Datasets and R scripts used for statistical analysis are available on https://github.com/francescochecchi/yem_burials_satellite_imagery. Our original study design was to infer countrywide mortality directly based on the sample of subdistricts. However, across subdistricts we were able to generate data for only a minority of cemeteries (see Results), restricting us to an exploration of burial patterns within this probably unrepresentative sample of remaining cemeteries.

First, we imputed missing grave count values (Table 1) by training a predictive generalised linear mixed model (GLMM) on longitudinal cemetery observations with complete surface area and grave count (Sq Appendix, Fig S12). The model estimated new graves since the previous image as a function of the natural log of new surface area with starting grave count as an offset, cemetery as a random effect and a negative binomial distributional assumption. We validated the model for prediction using leave-one-out cross-validation (S1 Appendix, Fig S13). For more detail, see Koum Besson et al. [17].

Next, we sought to model the number of new graves between each image observation, so as to explore either the association of burial with factors considered proxies of crisis exposure (incidence of insecurity events; price of wheat; proportion of IDPs in the population), adjusted for possible confounders; or the ratio of predicted burials under observed conditions versus under assumed counterfactual values of the crisis-proxy predictors. We assumed that counterfactual (i.e. no crisis) conditions would have consisted of zero insecurity events, zero displacement and the district-specific mean wheat price before the crisis period (March 2009, start of the price time series, to May 2014). We divided predictions under observed and counterfactual conditions to compute a burial rate ratio for each cemetery, as well as a crude overall rate ratio by summing predictions across all cemeteries.

We initially fit generalised linear or additive mixed models (with cemetery as random effect) to the counts of incident graves: both, however, featured extreme coefficient instability due to multi-collinearity, even after centring and standardising all continuous predictors. This restricted us to approaches that do cope with multicollinear data, but for which existing, well-documented statistical methods and packages have limited applicability (some do not account for longitudinal/grouped observations; others do not model count data). Specifically, we fitted the following alternatives: (i) a random forest (`ranger` package [30]) regression of the logged continuous burial rate (graves per day) with 1000 trees, minimum node size of 2 and weights for each observation corresponding to the proportion of all new graves in the sample accounted for by the cemetery to which the observation belonged; (ii) an elastic net generalised linear Poisson model (`glmnet` package [31]) of the counts of new graves, offset by the duration of each inter-observation period and weighted as above; this model, a compromise between ridge and LASSO regression, enables selection of a set of predictors even when multicollinearity is high, though it does not produce easily interpretable coefficients; and (iii) a Bayesian kernel regression machine (BKMR) model (`bkmr` [32] and `bkmrhat` packages) of the logged continuous burial rate, which treats exposures as an inherently correlated mixture of factors with a potentially hierarchical structure, while selecting out single exposures that do not contribute significantly to the outcome [33]. BKMR allows for grouped observations, adjustment variables and non-linear exposure-outcome relationships.

### Ethics statement

The study was approved by the Ethics Committee of the London School of Hygiene and Tropical Medicine (Reference: 22080). No data on live human participants were collected, and as such no consent was sought. All data has been previously collected for non-research purposes and the resolution of satellite imagery did not enable identification of people or other unique identifiers.

## Results

### Composition of the sample

#### Target sample

Altogether, the 24 sampled subdistricts contained an estimated 1,956,000 people (about 6% of Yemen’s total population of 30.9M as of September 2021; Table 2, Fig 3).

**Fig 3.**
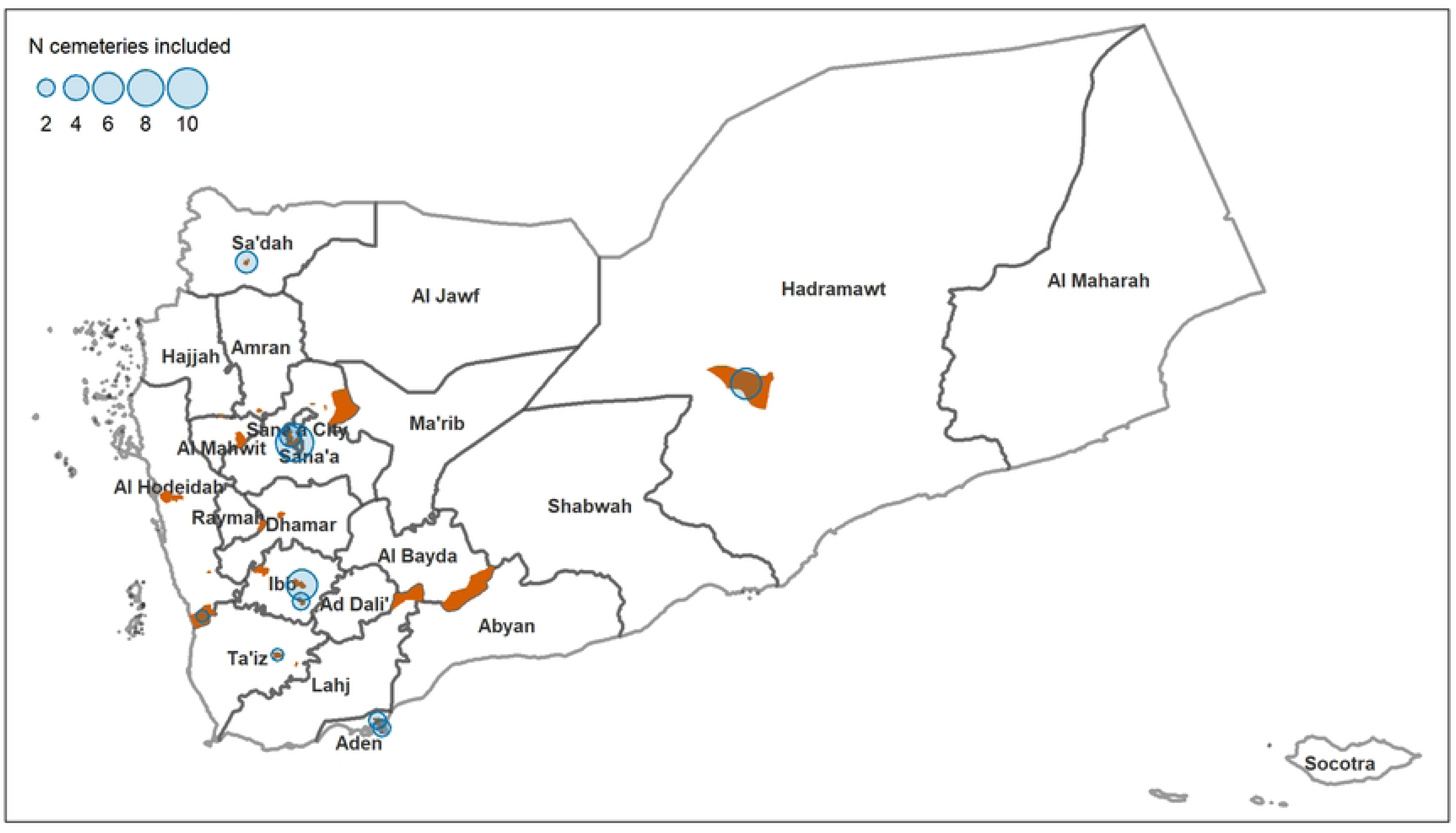
Map of Yemen governorates, with location of sampled subdistricts (orange-coloured polygons) and number of cemeteries included in analysis within each subdistrict (blue circles).

**Table 2.**
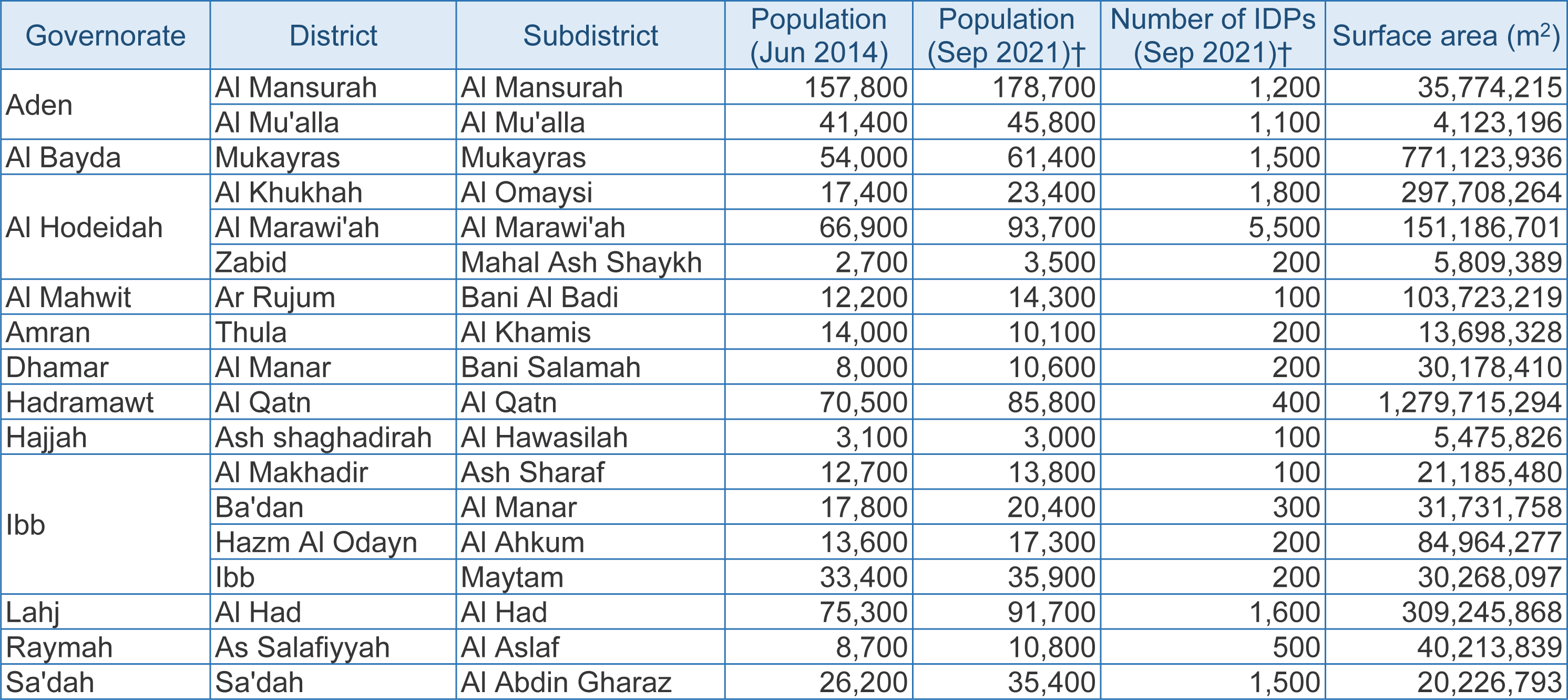
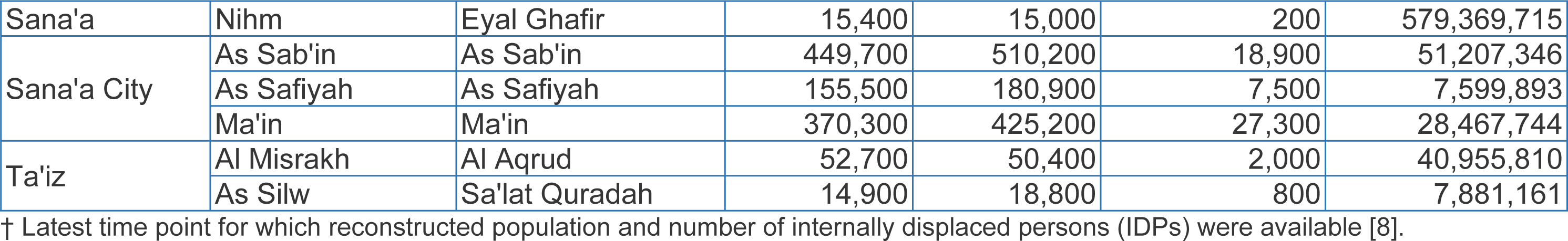
Characteristics of sampled subdistricts.

#### Sample attrition

The process of identifying possible cemeteries could not be completed in four subdistricts (Table 3). Out of 800 cemeteries initially listed, 239 were excluded for reasons not entailing potential bias (Fig 2). A further 467 were excluded at initial inspection as the cemetery was simply not locatable or visible, or appeared hard or impossible to analyse given available image quality even if some graves were visible. Notably, 205 cemeteries reviewed at this stage did have some visible graves, representing a minimum plausible denominator for the true number of cemeteries within the subdistricts. There were further data losses during geospatial analysis, largely because several cemeteries had < 2 images with robust data. Only 35 cemeteries in10/24 subdistricts were ultimately included in statistical analysis, namely 17.1% of those with any visible graves (Fig 2, Table 3, Fig 3). Further detail is provided in S1 Appendix, Table S1.

**Fig 2.**
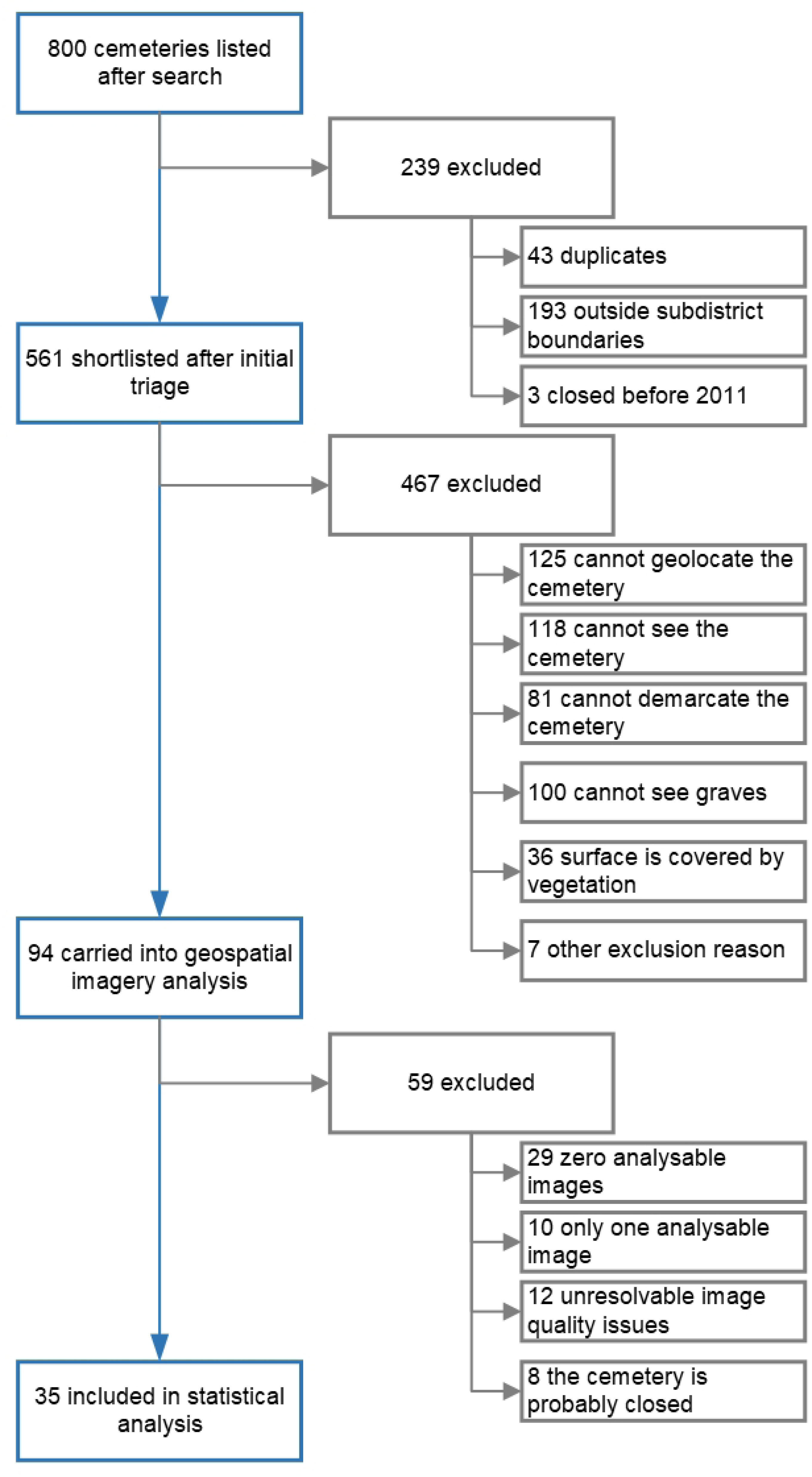
Flowchart of steps leading to final cemetery sample, with reasons for exclusion.

**Table 3.**
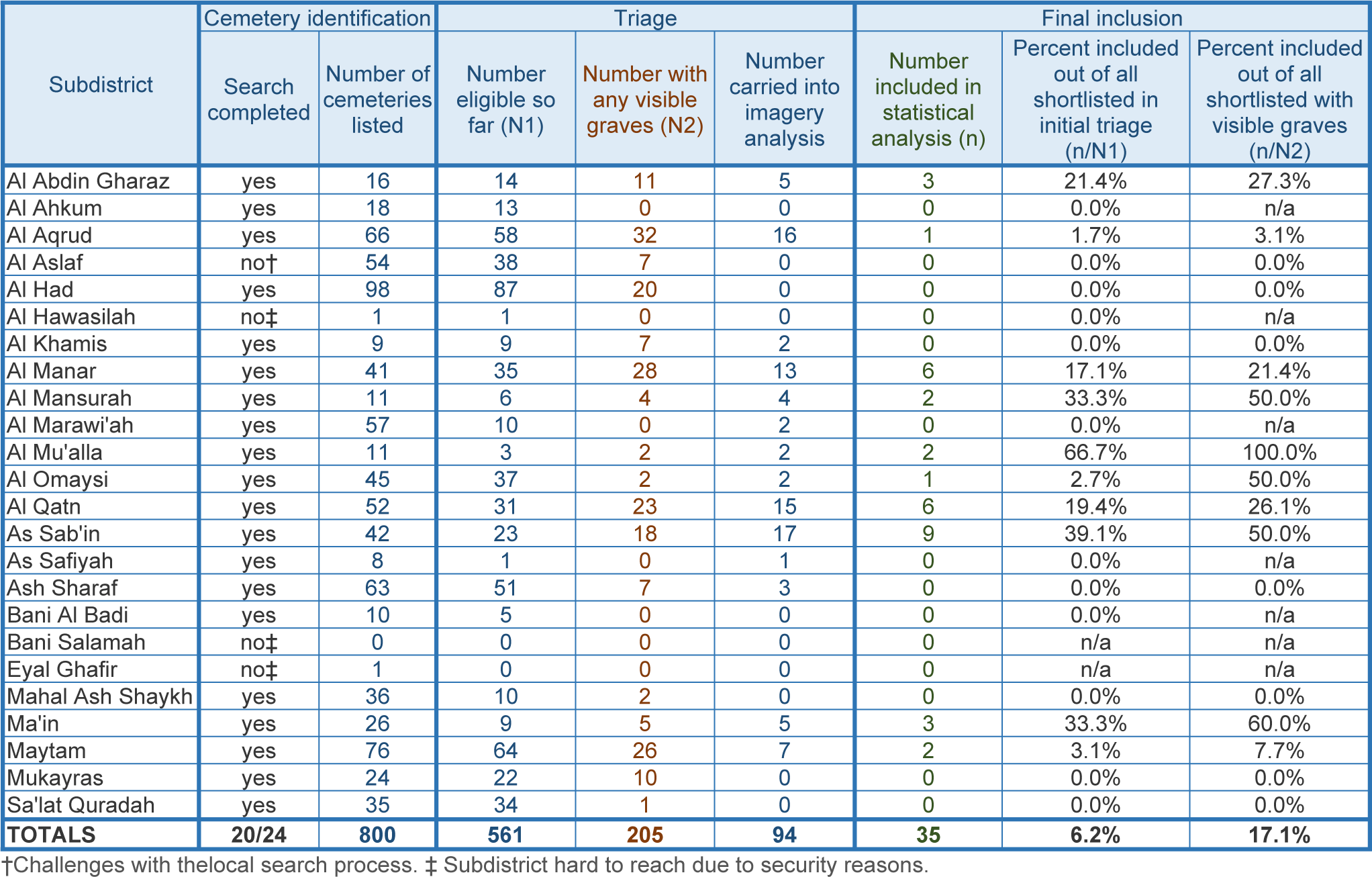
Composition of the final sample of cemeteries, by subdistrict.

### Characteristics of included cemeteries

Out of the 436 possible cemeteries that passed initial triage and could be geolocated, 18/35 (51.4%) of cemeteries included were located in an urban setting, compared to 23/378 (5.7%) of those excluded (Chi-square p < 0.001). Of cemeteries included, 27/35 (77.1%) were on sandy terrain, 4/35 (11.4%) were surrounded by vegetation and the remainder (4/35, 11.4%) on other terrain types; these proportions were 240/378 (59.9%), 115/378 (28.7%) and 46/378 (11.4%) among excluded cemeteries, respectively (Chi-square p = 0.059).

Of the 35 included cemeteries, 9 had two, 15 three, 10 four and one (previously analysed in the pilot Aden study) had seven analysable images, for a total of 110 images (mean = 3.1 per cemetery). Image availability was highest from mid-2018 onwards (Fig 4). The median starting number of graves was 317 (IQR 36 to 1110). Of 28 cemeteries with measurable surface area (i.e. excluding seven with a sparse layout: see Methods), the median starting area value was 3593m^2^ (interquartile range, IQR 1072m^2^ to 10,312m^2^).

**Fig 4.**
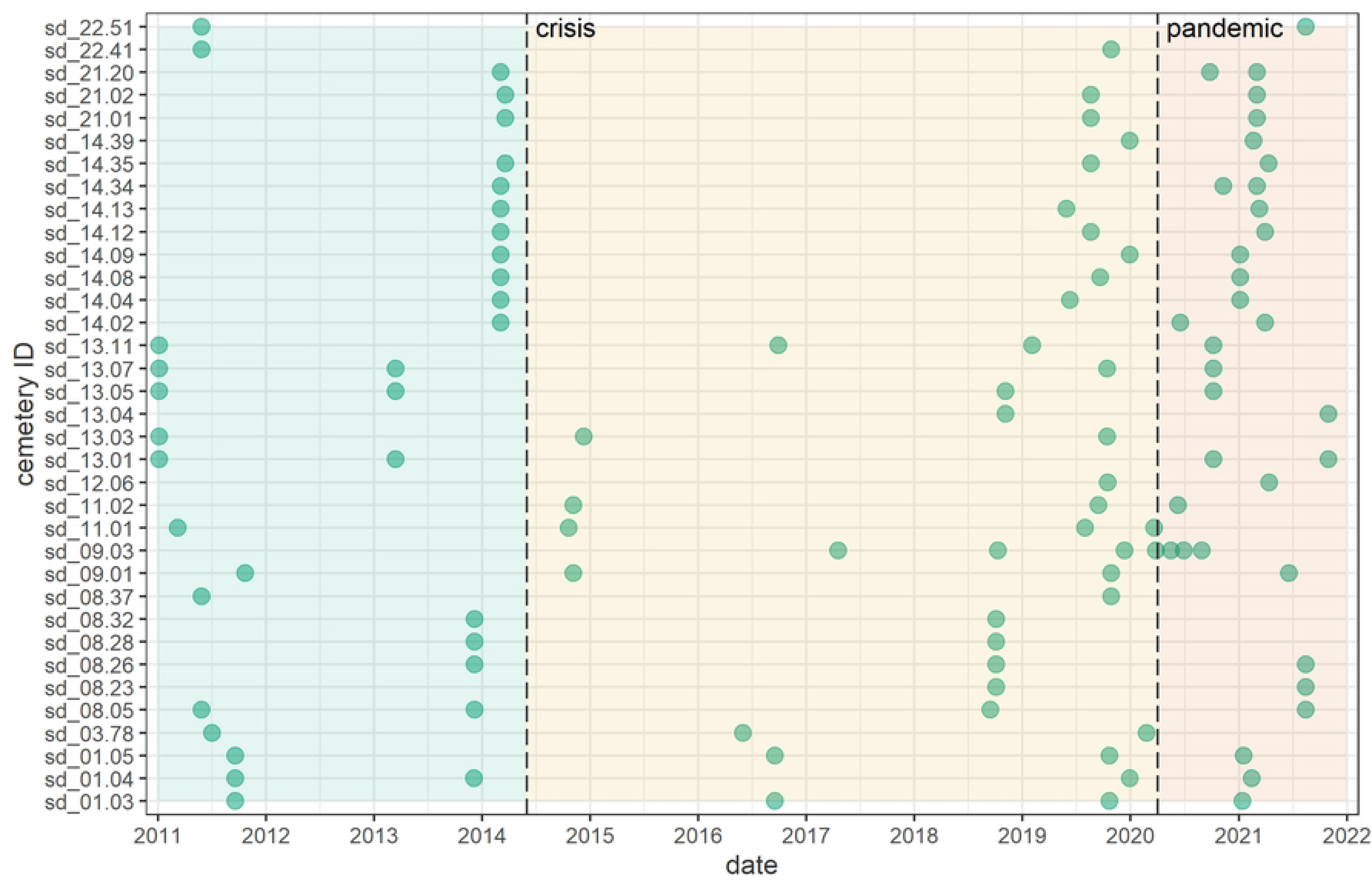
Dates on which analysable images (n = 110) were collected, by cemetery (N = 35).

### Burial rate patterns

Across the 35 cemeteries and 75 inter-image periods, the mean burial rate per day ranged from 0.00 to 2.57, with a median of 0.06 (IQR 0.02 to 0.39). When considering inter-image periods after the first in each cemetery, the mean burial rate tended to decrease during the years 2013 to 2018 and increase thereafter (note that the last periods overlap fully or partially with the pandemic; Fig 5). Burial rate by cemetery is shown in S1 Appendix, Fig S14.

**Fig 5.**
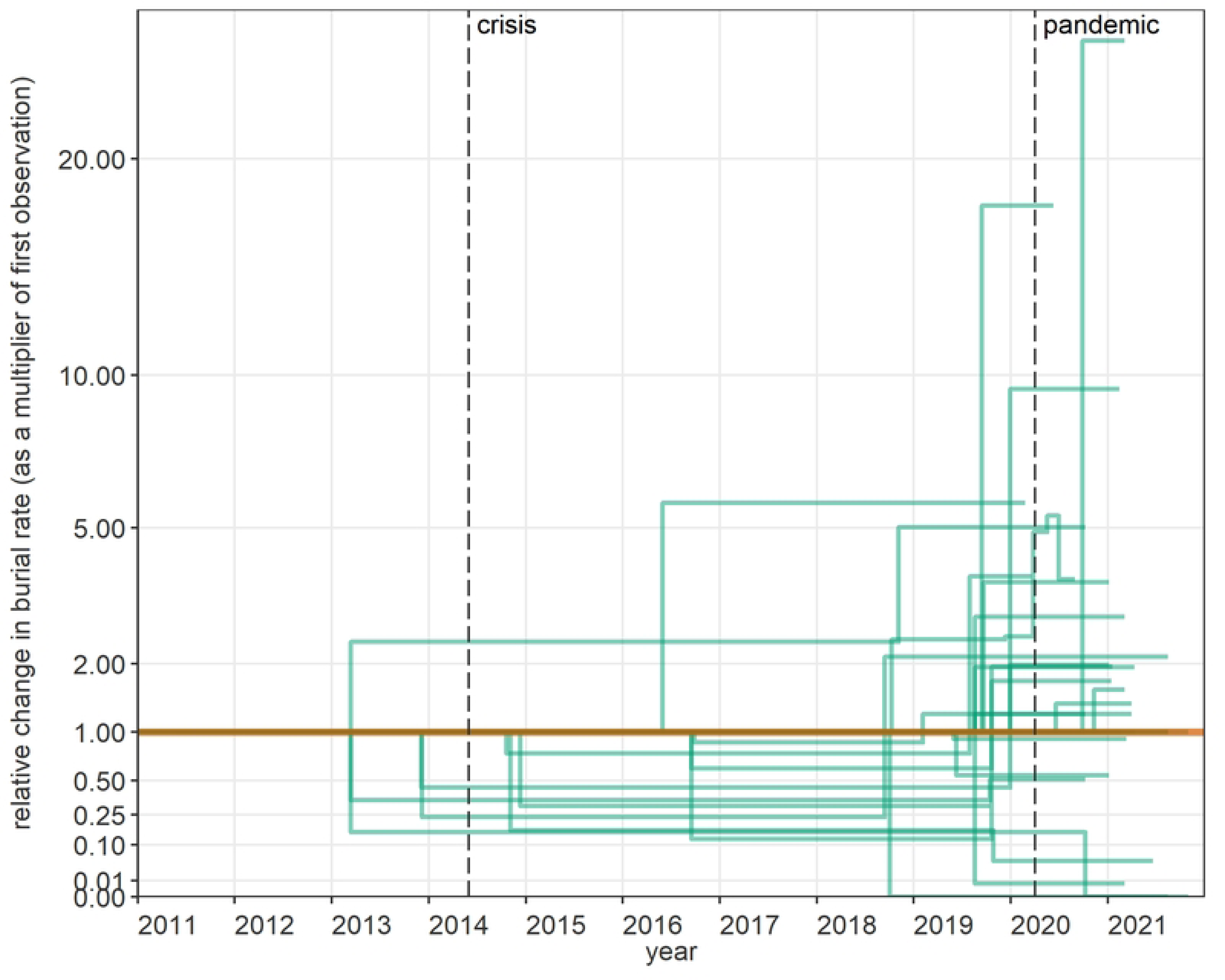
Relative change in burial rate from the first inter-image period in each cemetery time series. Each step function (green line) is one cemetery (all cemeteries start at a baseline of 1, coloured orange).

Of the three statistical models applied to the data, random forest yielded the best predictive accuracy despite not accounting for the longitudinal data structure (Fig 6), while the BKMR model was imprecise and featured considerable downward bias in the largest cemeteries (S1 Appendix, Fig S15). The random forest and elastic net model suggested that, under observed conditions, overall burials across our sample were about twice than they would have been if the three crisis proxies (insecurity, wheat price, displacement) had taken no-crisis counterfactual values, with most cemeteries experiencing elevated burials. However, the individual-cemetery predicted burial ratios were inconsistent between the two models (Fig 6). No confidence intervals were computed for these ratios, as no straightforward contrast or bootstrap method was found to divide random forest predicted distributions given the unknown correlation between predictions under observed and counterfactual conditions; moreover, the elastic net regression does not generate meaningful coefficient standard errors. Predictions by cemetery are shown in S1 Appendix (Fig S16, Fig S17, Fig S18, Fig S19).

**Fig 6.**
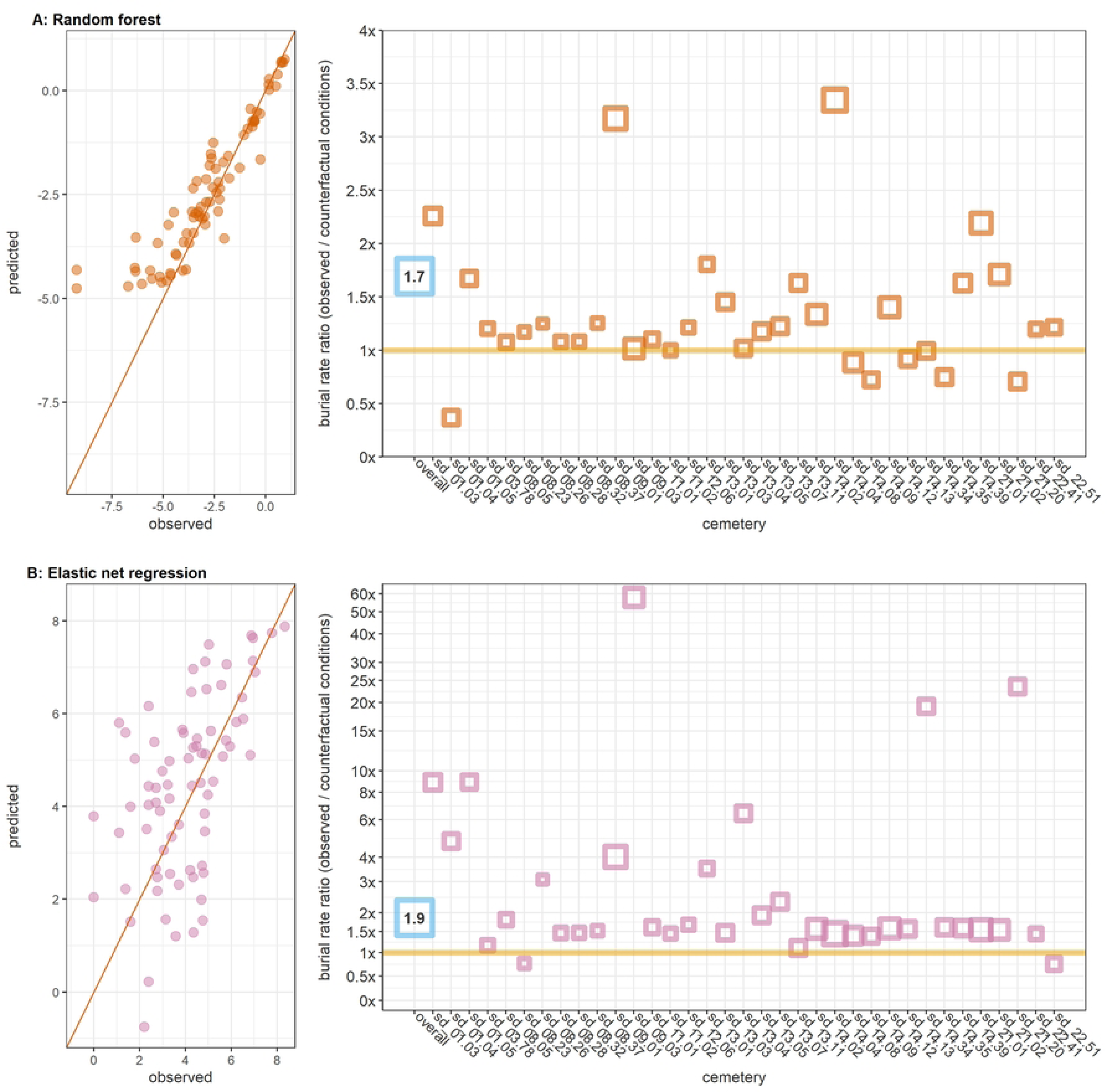
Results of the random forest (panel A) and elastic net (panel B) regression models. The leftmost graphs show model predictions versus observations (log burial rate for random forest, log incident burials for elastic net). The rightmost graphs show the ratio of cumulative burials predicted under observed versus counterfactual (no crisis) conditions for each cemetery and overall, with each square size proportional to the cemetery’s relative contribution to all burials within the sample: note the very different y-axis scales.

After fitting a BKMR model, the exposure-outcome relationship for each component of the overall exposure mixture can be visualised by holding the other components constant, as shown in Fig 7. This analysis suggests that the log burial rate increases linearly as a function of increasing insecurity events, with (counterintuitively) the opposite pattern for wheat price and no clear association for IDP proportion. The posterior probabilities of inclusion in the exposure mixture were 1.00, 0.59 and 0.41.

**Fig 7.**
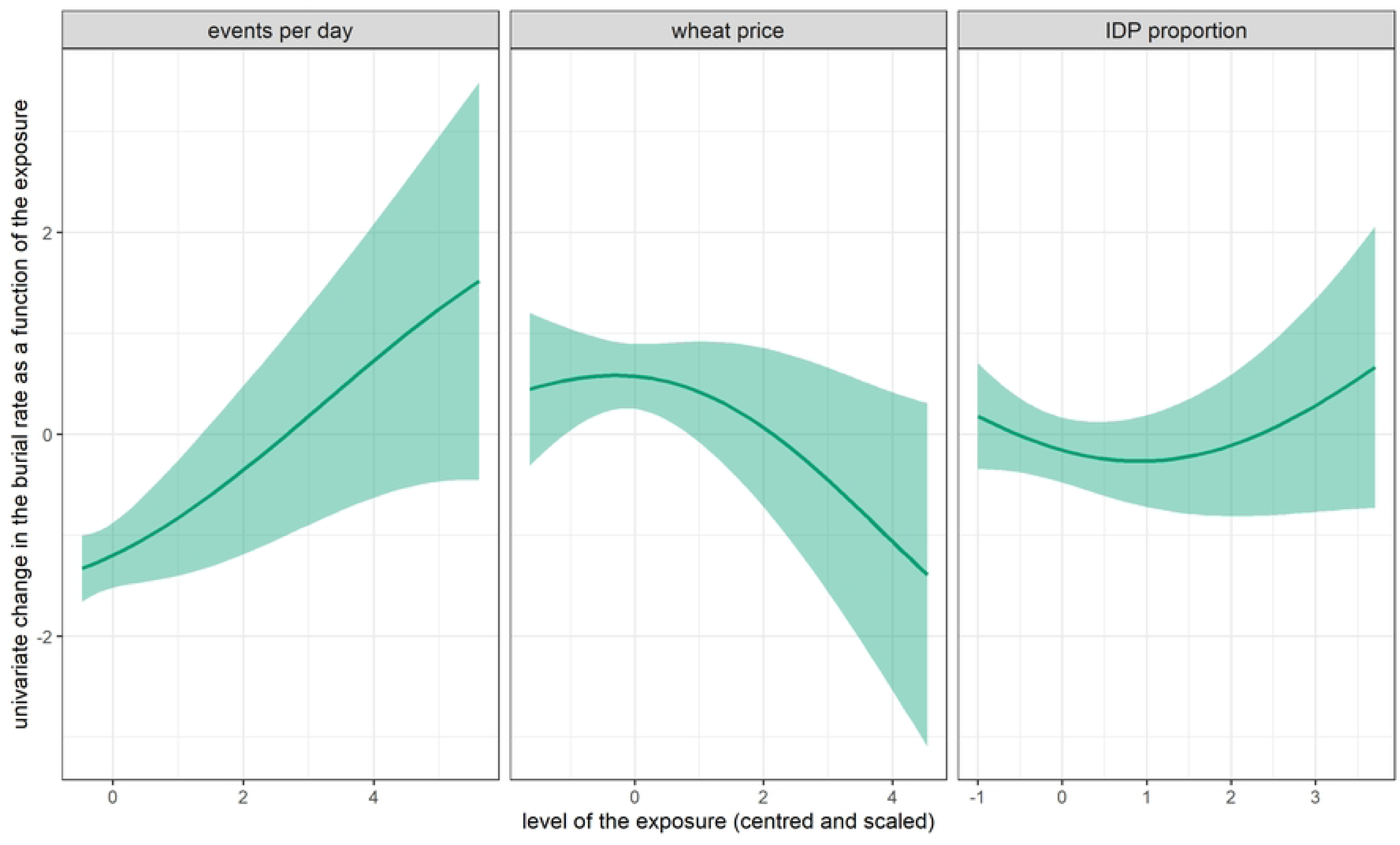
Relationship between each crisis exposure variable in the assumed mixture and the outcome (log burial rate), when holding the other exposures at their median values, and all other model covariates at their observed values, as estimated by Bayesian kernel mixture regression (BKMR). Shaded areas indicate 95% confidence intervals.

## Discussion

### Main findings

To our knowledge this is the largest-scale and one of the few published analyses of cemetery burial patterns as detectable from satellite imagery. While this data source may be of minimal utility in settings with accurate vital events registration, elsewhere, and especially in hard-to-access locations, it is potentially valuable to generate robust estimates of mortality and excess mortality due to crises or other public health threats, such as the COVID-19 pandemic, and thereby illuminate the true toll of these events in terms of its sheerest metric, namely human survival. Combinations of satellite imagery, ground observations and drone photography have been used recently to quantify old graves in the United States [34] and document potential war crimes in Syria [35] and Ukraine [36].

Contrary to a pilot study in Aden [17], but consistent with subsequent experience in Mogadishu, Somalia [20], this study presented numerous challenges with identification of cemeteries, quality image availability and geospatial analysis, which, taken together, impeded our original objective of countrywide, representative estimation. However, results suggest some broad patterns in terms of mortality during the crisis and pandemic periods in Yemen, and lay a foundation for further methods development. In our limited sample of 35 cemeteries, burial rate mostly decreased from baseline during the first years of the crisis, but increased sharply from 2019 onwards. Alternative models suggested a general pattern of increased burials compared to counterfactual scenarios in which key proxy variables for crisis conditions were held at the assumed no-crisis baseline. The incidence of insecurity events seemed to be associated with increasing burial rate, but the opposite was seen for the price of wheat.

### Limitations

Despite its nationally representative design, the study achieved a much-reduced sample, with evidence of selection bias towards urban locations. Rural locations with relatively small and not well-visible cemeteries, underrepresented in our sample, would plausibly have experienced higher mortality and thus burial. Therefore, inference about burial patterns across Yemen is severely limited. In addition, statistical results are potentially subject to random or systematic error due to inaccuracy in the model predictors used and in geospatial techniques to count graves and measure cemetery surface area. As findings do not feature confidence intervals, sampling variability and error in the imputation model for missing grave counts are not visualisable, impeding statistical significance assessment. The predictors and counterfactual assumptions used in the statistical model are limited and could have included additional variables that may be proxies of crisis conditions, such as climate.

While we attempted to reduce geospatial analysis error by omitting low-quality and otherwise problematic images from statistical analysis, some error is likely to have affected the final dataset. We assumed a planar terrain despite Yemen featuring many mountainous areas, which could have produced underestimation of surface area and problems with image orthorectification and alignment. Despite intensive expert supervision, the crowd worker approach carries a risk that serious analyst error may go unobserved: had resources been greater, this approach should have been validated more rigorously.

Generally, this unprecedented study highlights key challenges (Table 1) of analysing cemetery satellite images and identifying / counting graves across a variety of terrains and burial typologies given current imagery resolution. It is also possible that in locations with very high levels of violence, mass graves or less formal burials would have remained undetected by our analysis.

### Conclusions and ways forward

This analysis provides limited evidence on the possible excess in burials and thus population mortality attributable to crisis conditions in Yemen since 2014. It also documents a number of challenges that other analysts may encounter when attempting a similar analysis. In addition to some of the problems we describe above, the ethics of remote satellite imagery analysis, and particularly how to involve local actors in analyses without endangering their security, deserves careful consideration.

Based on experience to date, we believe that long-term monitoring of well-selected sites, particularly in urban settings where cemeteries are likely to be more visible and imagery cover greater, is a reasonable option for mortality estimation in settings where other methods are impracticable and the population universally uses cemeteries. Whether nationally or regionally representative estimates of burial trends are feasible using satellite imagery remains unclear. Acquisition of customised imagery is a service provided by satellite imagery companies and, while much more expensive than archive imagery, could greatly improve data quality. Furthermore, our experience suggests several avenues for methodological improvement. Estimating the proportion of graves likely to be missed based on image characteristics and quality and correcting grave counts accordingly, rather than omitting the image from analysis altogether, could greatly increase data quantity. Automated image quality metrics [37] could also be considered. The assumption of planar terrain could be relaxed by integrating elevation data (e.g. the Advanced Spaceborne Thermal Emission and Reflection Radiometer Global Digital Elevation Model [38]) in orthorectification or, more labour-intensively, placing a far greater number of tie-points in each consecutive image. The value of automated annotation of graves should be reviewed: on the one hand, asking human annotators to identify graves from scratch may be less confusing and more accurate; on the other hand, further validation of machine learning algorithms could greatly streamline analysis and reduce costs considerably, as suggested by a pilot analysis of cemeteries in North and South Korea [39]. Another cost-saving measure could be to invite citizen volunteers to perform annotation work. As regards study design, our experience suggests that purposeful selection of a few ‘sentinel sites’, with frequent observations per site, would be preferable to this study’s attempt at representative sample. Such sentinel time series could be combined with predictors, as in this analysis, to train and validate a model that predicts burial patterns indirectly, even where no imagery data are available. Satellite imagery analysis should not be a substitute for strengthening vital events registration and undertaking ground mortality estimation where possible, but does warrant further exploration and testing in a variety of settings.

## Data Availability

Datasets and R scripts used for statistical analysis are available on https://github.com/francescochecchi/yem_burials_satellite_imagery.

## Acknowledgments

The study was funded by the United Kingdom Foreign Commonwealth and Development Office through separate grants to the LSHTM (grant reference 300708-139) and the Satellite Applications Catapult (grant reference 300680-104). Funders were not involved in study design, data collection, analysis, or manuscript preparation.

We are grateful to Louise Mellor, Fergus McBean and other colleagues at the United Kingdom Foreign Commonwealth and Development Office for support. At the LSHTM, we are grateful to Lucy Bell and Catherine McGowan for project management support. The Satellite Applications Catapult would like to thank all the staff at 1715 Labs for their support throughout this project. We also acknowledge numerous Yemeni geographers and civil society members who helped our team identify potential cemeteries, including Odai Al-Masni, who fulfils authorship criteria but could not be reached when this paper was being drafted.

**S1 Appendix. Additional figures and tables.**

## References

1. United Nations Office for Coordination of Humanitarian Affairs. Yemen Situation Report. OCHA. 2023. https://www.unocha.org/yemen. Accessed 10 Feb 2023.

2. Maxwell D, Hailey P, Spainhour Baker L, Kim JJ. Constraints and Complexities of Information and Analysis in Humanitarian Emergencies: Evidence from Yemen. Feinstein International Center, Tufts University and Centre for Humanitarian Change; 2019.

3. Armed Conflict Location & Event Data Project (ACLED). Research Hub: War in Yemen. 2023. https://acleddata.com/research-hub-war-in-yemen/. Accessed 10 Feb 2023.

4. Dureab F, Al-Falahi E, Ismail O, Al-Marhali L, Al Jawaldeh A, Nuri NN, et al. An Overview on Acute Malnutrition and Food Insecurity among Children during the Conflict in Yemen. Children. 2019;6:77.

5. Hashim HT, Miranda AV, Babar MS, Essar MY, Hussain H, Ahmad S, et al. Yemen’s triple emergency: Food crisis amid a civil war and COVID-19 pandemic. Public Health in Practice. 2021;2:100082.

6. AlKarim T, Abbara A, Attal B. Armed conflict alone does not explain the devastation of Yemen’s health system. BMJ Glob Health. 2021;6:e004740.

7. Alsabri M, Alsakkaf L, Alhadheri A, Cole J, Burkle Jr. F. Chronic Health Crises and Emergency Medicine in War-torn Yemen, Exacerbated by the COVID-19 Pandemic. WestJEM. 2022;23:276–84.

8. Checchi F, Koum Besson ES. Reconstructing subdistrict-level population denominators in Yemen after six years of armed conflict and forced displacement. Journal of Migration and Health. 2022;5:100105.

9. Camacho A, Bouhenia M, Alyusfi R, Alkohlani A, Naji MAM, de Radiguès X, et al. Cholera epidemic in Yemen, 2016–18: an analysis of surveillance data. The Lancet Global Health. 2018;6:e680–90.

10. Alsabri M, Alhadheri A, Alsakkaf LM, Cole J. Conflict and COVID-19 in Yemen: beyond the humanitarian crisis. Global Health. 2021;17:83.

11. Rahmat ZS, Islam Z, Mohanan P, Kokash DM, Essar MY, Hasan MM, et al. Food Insecurity during COVID-19 in Yemen. The American Journal of Tropical Medicine and Hygiene. 2022;106:1589–92.

12. Kotiso M, Qirbi N, Al-Shabi K, Vuolo E, Al-Waleedi A, Naiene J, et al. Impact of the COVID-19 pandemic on the utilisation of health services at public hospitals in Yemen: a retrospective comparative study. BMJ Open. 2022;12:e047868.

13. Mikkelsen L, Phillips DE, AbouZahr C, Setel PW, de Savigny D, Lozano R, et al. A global assessment of civil registration and vital statistics systems: monitoring data quality and progress. The Lancet. 2015;386:1395–406.

14. Brolan CE, Gouda H. Civil Registration and Vital Statistics, Emergencies, and International Law: Understanding the Intersection. Medical Law Review. 2017;25:314–39.

15. Assesing the Impact of War on Development in Yemen | United Nations Development Programme. UNDP. https://www.undp.org/yemen/publications/assesing-impact-war-development-yemen. Accessed 10 Feb 2023.

16. Alhaffar M, Basaleem H, Othman F, Alsakkaf K, Naji SMM, Kolaise H, et al. Adult mortality before and during the first wave of COVID-19 pandemic in nine communities of Yemen: a key informant study. Confl Health. 2022;16:63.

17. Koum Besson ES, Norris A, Bin Ghouth AS, Freemantle T, Alhaffar M, Vazquez Y, et al. Excess mortality during the COVID-19 pandemic: a geospatial and statistical analysis in Aden governorate, Yemen. BMJ Glob Health. 2021;6:e004564.

18. COVID-19 pandemic in Yemen. Wikipedia. 2022.

19. Raleigh C, Linke A, Hegre H, Karlsen J. Introducing ACLED-Armed Conflict Location and Event Data. Journal of Peace Research. 2010;47:1–10.

20. Warsame A, Bashiir F, Freemantle T, Williams C, Vazquez Y, Reeve C, et al. Excess mortality during the COVID-19 pandemic: a geospatial and statistical analysis in Mogadishu, Somalia. International Journal of Infectious Diseases. 2021;113:190–9.

21. Yemen Central Statistical Organisation. Yemen - Roads. Humanitarian Data Exchange. 2018. https://data.humdata.org/dataset/yemen-roads. Accessed 16 Dec 2021.

22. Humanitarian OpenStreetMap Team. Yemen Health Facilities (OpenStreetMap Export). Humanitarian Data Exchange. 2020. https://data.humdata.org/dataset/hotosm_yem_health_facilities. Accessed 16 Dec 2021.

23. Ministry of Public Health and Population - MOPHP/Yemen, Central Statistical Organization - CSO/Yemen, Pan Arab Program for Family Health - PAPFAM, ICF International. Yemen National Health and Demographic Survey 2013. Rockville, Maryland, USA: MOPHP, CSO, PAPFAM, and ICF International; 2015.

24. United Nations World Food Programme. Yemen - Food Prices - Humanitarian Data Exchange. https://data.humdata.org/dataset/wfp-food-prices-for-yemen. Accessed 10 Feb 2023.

25. Vidhya GR, Ramesh H. Effectiveness of Contrast Limited Adaptive Histogram Equalization Technique on Multispectral Satellite Imagery. In: Proceedings of the International Conference on Video and Image Processing. Singapore Singapore: ACM; 2017. p. 234–9.

26. Mustafa WA, Abdul Kader MMM. A Review of Histogram Equalization Techniques in Image Enhancement Application. J Phys: Conf Ser. 2018;1019:012026.

27. Unsharp masking. Wikipedia. 2022.

28. NW 1615 L. St, Washington S 800, Inquiries D 20036 U-419-4300 | M-857-8562 | F-419-4372 | M. 4. Turkers in this canvassing: young, well-educated and frequent users. Pew Research Center: Internet, Science & Tech. 2016. https://www.pewresearch.org/internet/2016/07/11/turkers-in-this-canvassing-young-well-educated-and-frequent-users/. Accessed 10 Feb 2023.

29. Brodzik MJ, Billingsley B, Haran T, Raup B, Savoie MH. EASE-Grid 2.0: Incremental but Significant Improvements for Earth-Gridded Data Sets. IJGI. 2012;1:32–45.

30. Wright MN, Ziegler A. ranger: A Fast Implementation of Random Forests for High Dimensional Data in C++ and R. Journal of Statistical Software. 2017;77:1–17.

31. Friedman J, Hastie T, Tibshirani R. Regularization Paths for Generalized Linear Models via Coordinate Descent. J Stat Soft. 2010;33.

32. Bobb JF, Valeri L, Claus Henn B, Christiani DC, Wright RO, Mazumdar M, et al. Bayesian kernel machine regression for estimating the health effects of multi-pollutant mixtures. Biostatistics. 2015;16:493–508.

33. Bobb JF, Claus Henn B, Valeri L, Coull BA. Statistical software for analyzing the health effects of multiple concurrent exposures via Bayesian kernel machine regression. Environ Health. 2018;17:67.

34. Spera SA, Franklin MS, Zizzamia EA, Smith RK. Recovering a Black Cemetery: Automated Mapping of Hidden Gravesites Using an sUAV and GIS in East End Cemetery, Richmond, VA. Int J Histor Archaeol. 2022;26:1110–31.

35. Investigating Mass Graves using Satellite Imagery. Syria Justice & Accountability Centre. 2022. https://syriaaccountability.org/mass-graves-using-satellite-imagery/. Accessed 10 Feb 2023.

36. El-Sherbiny E, den Braber B. Mass graves after the Russian invasion: Bucha, Mariupol, Chernihiv, Kherson. London: Centre for Information Resilience; 2022.

37. CPBD | Image, Video, and Usability Lab. https://ivulab.asu.edu/software/cpbd/. Accessed 10 Feb 2023.

38. Abrams M, Crippen R, Fujisada H. ASTER Global Digital Elevation Model (GDEM) and ASTER Global Water Body Dataset (ASTWBD). Remote Sensing. 2020;12:1156.

39. Lunga D, Dhamdhere R, Walters S, Bragg L, Makkar N, Urban M. Learning to Count Grave Sites for Cemetery Observation Models With Satellite Imagery. IEEE Geosci Remote Sensing Lett. 2022;19:1–5.

